# Expanding HIV Clinical Monitoring: The Role of CD4, CD8, and CD4/CD8 Ratio in Predicting Non-AIDS Events

**DOI:** 10.1101/2023.03.31.23288001

**Authors:** Javier Martínez-Sanz, Jorge Díaz Álvarez, Marta Rosas, Raquel Ron, José Antonio Iribarren, Enrique Bernal, Félix Gutiérrez, Federico García, Noemi Cabello, Julián Olalla, Santiago Moreno, Sergio Serrano-Villar, CoRIS

## Abstract

**Background:** While a low CD4/CD8 ratio during HIV treatment correlates with immunosenescence, its value in identifying patients at an increased risk for clinical events remains unclear.

**Methods:** We analyzed data from the CoRIS cohort to determine whether CD4 count, CD8 count, and CD4/CD8 ratio at year two of antiretroviral therapy (ART) could predict the risk of serious non-AIDS events (SNAEs) during the next five years. These included major adverse cardiovascular events, non-AIDS-defining malignancies, and non-accidental deaths. We used pooled logistic regression with inverse probability weighting to estimate the survival curves and cumulative risk of clinical events.

**Results:** The study included 4625 participants, of whom 4.3% experienced an SNAE during the follow-up period. A CD4/CD8 ratio <0.3 predicted an increased risk of SNAEs during the next five years (OR 1.63, 95%CI 1.03-2.58). The effect was stronger at a CD4/CD8 ratio cut-off of <0.2 (OR 3.09, 95%CI 1.57-6.07). Additionally, low CD4 count at cut-offs of <500 cells/μL predicted an increased risk of clinical events. Among participants with a CD4 count ≥500 cells/μL, a CD8 count ≥1500 cells/μL or a CD4/CD8 ratio <0.4 predicted increased SNAE risk.

**Conclusions:** Our results support the use of the CD4/CD8 ratio and CD8 count as predictors of clinical progression. Patients with CD4/CD8 ratio <0.3 or CD8 count ≥1500/μL, regardless of their CD4 count, may benefit from closer monitoring and targeted preventive interventions.

**Summary:** This study found that a low CD4/CD8 ratio (<0.3) or a CD8 count ≥1500/μL after two years of antiretroviral therapy predicts an increased risk of serious non-AIDS events, regardless of CD4 count. These patients may benefit from closer follow-up.

## Background

People living with HIV (PWH) continue to have a higher risk of mortality and severe clinical events than people without HIV, despite the efficacy of current antiretroviral therapy (ART) [1–3]. In this population, CD4 count normalization does not reflect a complete return-to-health state. Persistent residual immune dysfunction contributes to an increased risk of severe non-AIDS events (SNAEs) [4,5].

The CD4/CD8 ratio has emerged as a useful indicator of immune dysfunction in PWH. It can be easily monitored in routine clinical practice and correlates with markers of immunosenescence and inflammation, which are technically difficult to measure and too variable to be used at an individual level [6,7]. PWH with a low CD4/CD8 ratio exhibit increased inflammation and immunosenescence despite successful ART (i.e., having achieved a CD4 count >500 cells/μL) [8].

Several studies have investigated whether the CD4/CD8 ratio predicts the risk of SNAEs or mortality in PWH. However, results are conflicting, with variations in study designs, timing of ratio measurement, ratio thresholds, and endpoints. While some studies have found a significant association between a low CD4/CD8 ratio and an increased risk of SNAEs or mortality [7–12], others have failed to find this association [14,15]. This lack of consensus has led to discrepancies in clinical guidelines regarding the appropriateness of monitoring the CD4/CD8 ratio [16,17]. Most evidence linking this marker to clinical outcomes has been generated in retrospective cohort studies, which may be limited by the lack of data on CD8 count, high rates of loss to follow-up, and inability to adjudicate clinical events [8,9,13,15,18]. The most informative cut-off values are still unclear, as are the type of events that these markers can predict, and the timing of the measurement relative to ART initiation [8,9,12–14,19]. Furthermore, extracting the independent impact of CD8 count is difficult, as their increase may be a homeostatic response to a low CD4 count [19].

In this study, we aimed to assess whether the CD4/CD8 ratio and CD8 count provide additional prognostic information to CD4 count when measured at year two of ART and to determine the most discriminative thresholds in a large prospective cohort of PWH with long-term follow-up.

## Methods

### Study population

We used data from CoRIS, a prospective multicenter cohort of treatment-naïve adults with HIV, with standardized data collection since 2004. CoRIS collects data from 45 Spanish hospitals. In addition to demographic and clinical data, blood samples are collected and stored in a centralized biobank for the entire cohort. A baseline sample is collected before ART initiation and a follow-up sample is collected annually thereafter. Internal quality controls are performed annually, and 10% of the data are externally audited every two years [20]. We included individuals with ART initiation up to December 2014 (to allow for a 7-year follow-up) and HIV-RNA <50 copies/mL after two years of ART. We excluded participants with a history of AIDS events or SNAEs, and those with a CD4/CD8 ratio not measured at year two of ART (±3 months).

### Prognostic variables, follow-up, and outcomes

We set the index date (baseline) as the visit occurring two years (± 3 months) after ART initiation. Follow-up ended at the earliest loss to follow-up (last date with data update in CoRIS), ART discontinuation for more than 28 days, or administrative end of follow-up.

We explored different prognostic variables at baseline, including (i) reaching a CD4 count above 200, 350, and 500 cells/μL and (ii) reaching a CD4/CD8 ratio above the cut-off values of 0.2, 0.3, 0.4, and 0.5. These thresholds were selected before analysis and were based on previous studies [8,9,12–14]. Furthermore, we assessed the ability of CD8+ count (at 800, 1000, and 1500 cells/μL cut-offs) [14,19] and CD4/CD8 ratio to predict the outcome in the subpopulation of participants with a CD4+ count > 500 cells/μL at year two.

The primary outcome was the cumulative incidence of the SNAEs (major adverse cardiovascular event -MACE-, non-AIDS-defining malignancy -NADM-, or non-accidental death as a composite event) during the subsequent five years. The secondary outcomes included the cumulative incidence of (i) MACE, (ii) NADM, and (iii) nonaccidental death. MACE was a composite of nonfatal myocardial infarction, nonfatal stroke, and cardiovascular death.

### Statistical methods

We estimated the five-year risk of experiencing a clinical event using double-weighted pooled logistic regression. Additionally, we computed survival curves by introducing a time-varying intercept to allow the hazard to vary over time. We used inverse probability weighting to control for confounding variables. Furthermore, to address informative censoring, we applied inverse probability of censoring weighting (IPCW) for participants who did not remain in follow-up through year five. Participants accumulated weights until their event or the end of follow-up, whichever occurred first. The estimated weights were truncated at their 99^th^ percentile to prevent outliers from affecting the analyses. The IPCW maintains a constant effective sample size (N = 4,625 at ART year 2) for each year of follow-up, transferring the statistical weight of lost or censored participants to those remaining under observation [21].

Covariates included in the pooled logistic regression were age, sex, date of enrolment, mode of HIV transmission, education level, geographical origin, AIDS diagnosis, HIV RNA at diagnosis, type of ART regimen used (a boosted protease inhibitor regimen, a non-nucleoside reverse transcriptase inhibitor, or an integrase inhibitor), and nadir CD4 count. We also included CD8 count at baseline as a covariate in the models for CD4 count. To assess whether the prediction varied for different types of events, we separately conducted analyses on MACE, NADM, and non-accidental death. In a secondary analysis, we performed an *on-treatment* analysis. We assumed that patients who discontinued ART might have remained on treatment had they known of the potential adverse consequences of discontinuing ART. We performed all analyses using Stata v.17 (StataCorp LP, College Station, TX, USA).

## Results

We included 4625 participants. The study flowchart is shown in **Figure S1**. The median age was 37 years, 87% were male, and the median CD4/CD8 ratio at ART initiation was 0.29 (p27-p75 0.17, 0.46). Participants who had an event during follow-up showed a higher percentage of intravenous drug use as a risk factor for HIV transmission, lower educational level, lower nadir CD4 count, and lower CD4/CD8 ratio (**Table 1**).

**Table 1.** Baseline characteristics of the study population.

| **Baseline\* characteristics** | **Event (n = 200)** | **No event (n = 4,425)** |
| --- | --- | --- |
| **Age, median (p25, p75)** | 47 (39, 54) | 37 (30, 43) |
| **Gender, male, n (%)** | 165 (83) | 3687 (83) |
| **Mode of transmission, n (%)** |  |  |
| MSM | 75 (38) | 2534 (57) |
| Heterosexual | 36 (18) | 355 (8) |
| IDU | 81 (41) | 1396 (32) |
| Other | 8 (4) | 140 (3) |
| **Education level, n (%)** |  |  |
| None – Primary | 49 (25) | 658 (15) |
| Secondary – High school | 92 (36) | 2085 (47) |
| University | 28 (14) | 99 (22) |
| **Origin, n (%)** |  |  |
| Spain | 137 (69) | 2554 (58) |
| Latin America | 23 (11) | 726 (16) |
| Western Europe | 32 (16) | 773 (17) |
| Africa | 8 (4) | 245 (6) |
| Other | 0 (0) | 25 (1) |
| **AIDS diagnosis, n (%)** | 66 (33) | 790 (18) |
| **Nadir CD4+ (cells/μL), median (p25, p75)** | 173 (56, 264) | 250 (135, 342) |
| **CD4+ (cells/μL), median (p25, p75)** | 424 (266, 663) | 569 (404, 761) |
| **CD8+ (cells/μL), median (p25, p75)** | 880 (621, 1362) | 891 (661, 1198) |
| **CD4/CD8 ratio** | 0.48 (0.28, 0.81) | 0.64 (0.42, 0.92) |
| **ART regimen** |  |  |
| NNRTI | 92 (46) | 2631 (60) |
| PI | 90 (45) | 1408 (31) |
| INSTI | 9 (5) | 248 (6) |

*Abbreviations: ART, antiretroviral therapy; IDU, injecting drug use; INSTI, integrase strand transfer inhibitor; MSM, men who have sex with men; NNRTI, non-nucleoside reverse transcriptase inhibitor; PI, protease inhibitor.*
*\*Baseline was the visit at month 24 after ART initiation.*

Two years after ART initiation, 78%, 71%, 63%, and 53% of the participants had a CD4/CD8 ratio ≥0.2, ≥0.3, ≥0.4, and ≥ 0.5, respectively. Similarly, 78%, 67%, 50%, and 21% reached a CD4 count ≥ 200, 350, 500, and 750 cells/μL. Twelve percent were censored because of loss to follow-up (9%) or ART discontinuation (3%).

During the five-year follow-up, 4.3% had a SNAE: 48 (1%) had a MACE, 105 (2.3%) were diagnosed with a NADM, and 47 (1%) had non-accidental death, as shown in **Table 2**.

**Table 2.** Outcomes during follow-up.

| **Event** | **N (%)** |
| --- | --- |
| **Nonaccidental death** | **47 (23.5)** |
| **MACE** | **48 (24)** |
| Myocardial infarction | 35 (17.5) |
| Stroke | 13 (6.5) |
| **NADM** | **105 (52.5)** |
| Anus | 19 (9.5) |
| Hodgkin's lymphoma | 12 (6) |
| Skin (non-melanoma) | 10 (5) |
| Lung | 8 (4) |
| Liver | 7 (3.5) |
| Colorectal | 6 (3) |
| Non-specified metastasis | 6 (3) |
| Cervical | 5 (2.5) |
| Hematologic | 5 (2.5) |
| Prostate | 5 (2.5) |
| Soft tissue | 4 (2) |
| Head and neck | 4 (2) |
| Bladder | 3 (1.5) |
| Melanoma | 2 (1) |
| Uterus | 2 (1) |
| Breast | 1 (0.5) |
| Brain | 1 (0.5) |
| Pancreas | 1 (0.5) |
| Penis | 1 (0.5) |
| Stomach | 1 (0.5) |
| Testicle | 1 (0.5) |
| Vulva | 1 (0.5) |

*Abbreviations: MACE, major adverse cardiovascular event; NADM, non-AIDS-defining malignancy*

**Figure 1.** shows the survival curves and odds ratios (OR) for the event during follow-up for the CD4/CD8 models. A CD4/CD8 ratio <0.3 at year two of ART predicted an increased risk of SNAEs during follow-up (OR 1.63 [95% CI 1.03, 2.58]). At the lowest CD4/CD8 ratio cut-off (<0.2), the effect became stronger (OR 3.09 [95% CI 1.57, 6.07]). **Figure 2** shows the effect of CD4 count at year two on SNAE risk. A low CD4 count was associated with an increased risk of clinical events at cut-offs of 200, 350, and 500 cells/μL, above which the predictive ability was lost. Among participants with CD4 count ≥500 cells/μL, we found an increased risk of SNAEs at the 0.3 and 0.4 CD4/CD8 cut-offs (**Figure 3**). In analyses exploring the CD8 count effects in participants with a CD4 count ≥500 cells/μL, we found that a CD8 count ≥1500 CD8 cells/μL, but not the 800 and 1000 cells/μL cut-offs, predicted an increased risk of subsequent SNAEs (**Figure S2**).

**Figure 1.**
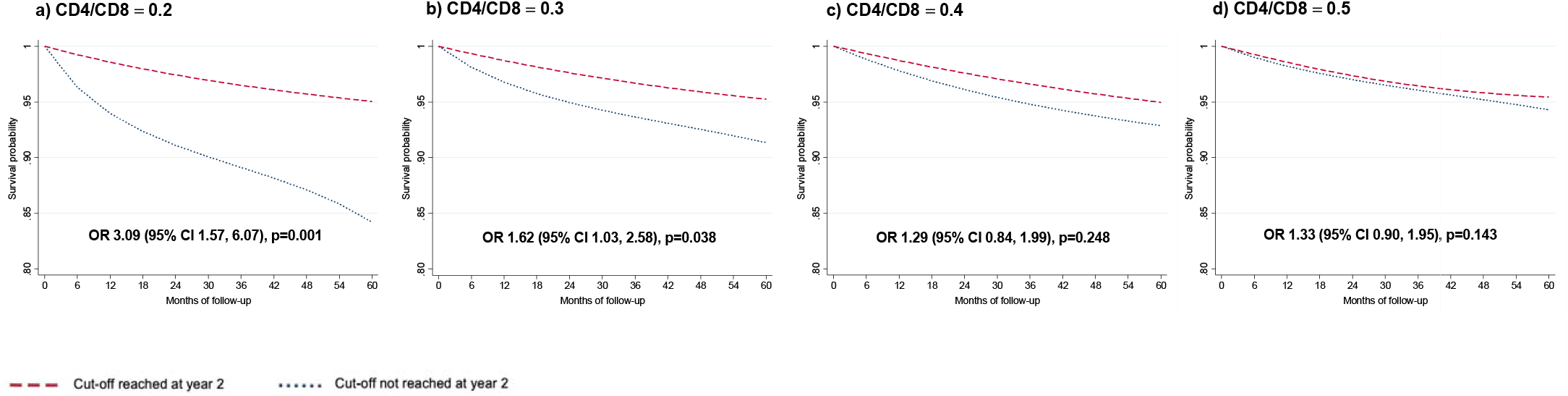
Survival curves for each CD4/CD8 cut-off. Survival probability and odds ratio (95% CI) for each subgroup of participants. The baseline visit (month 0 of follow-up) corresponds to 24 months after antiretroviral therapy initiation. The OR of presenting a clinical event corresponds to the five-year follow-up period.

**Figure 2.**
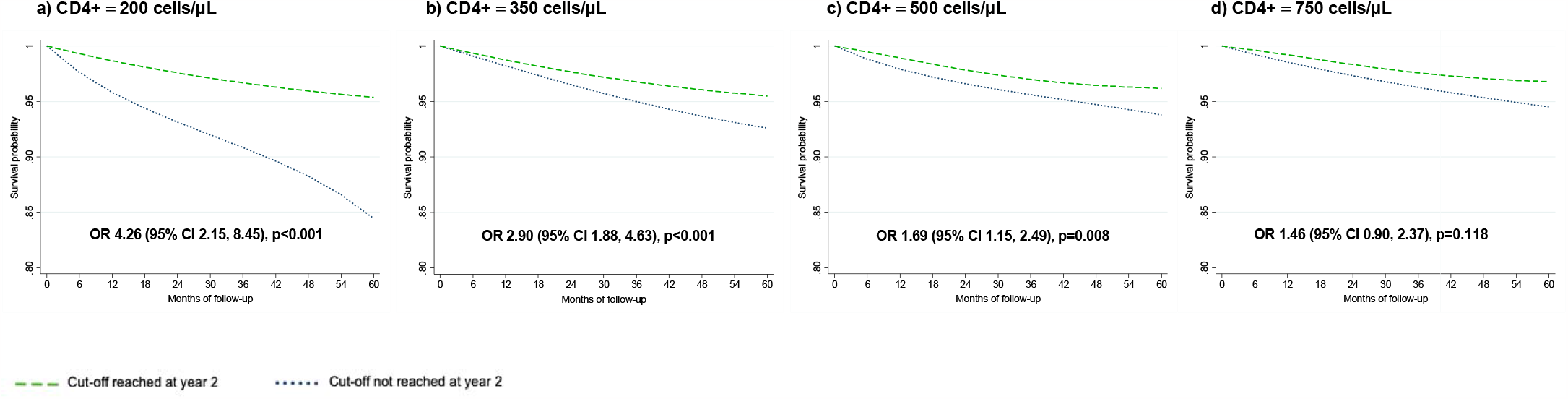
Survival curves for each CD4+ cut-off. Survival probability and odds ratio (95% CI) for each subgroup of participants. The baseline visit (month 0 of follow-up) corresponds to 24 months after antiretroviral therapy initiation. The OR of presenting a clinical event corresponds to the five-year follow-up period.

**Figure 3.**
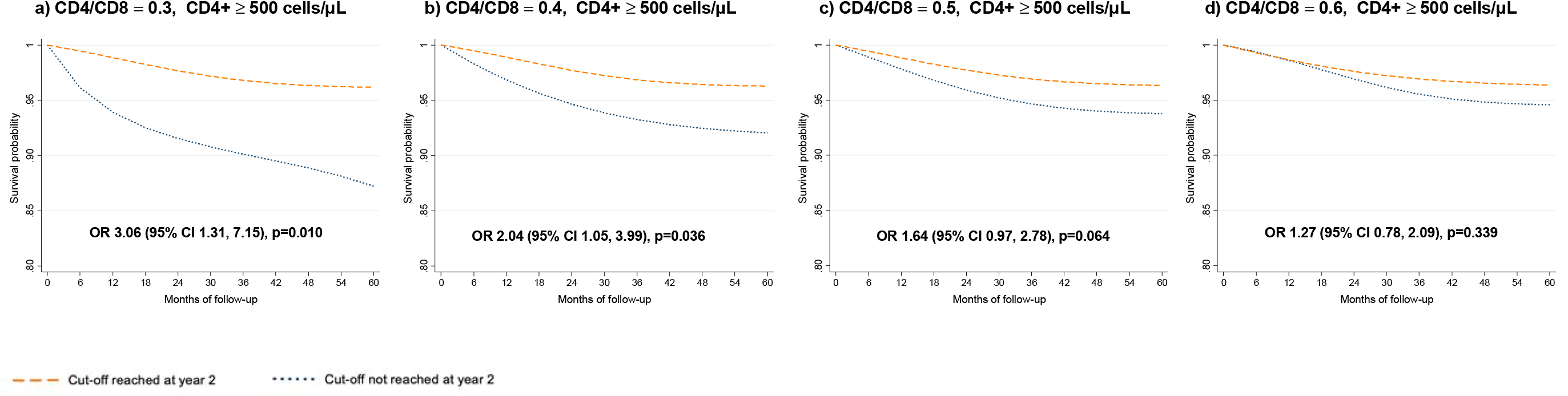
Survival curves for each CD4/CD8 cut-off among participants with CD4+ count ≥ 500 cells/μL. Survival probability and odds ratio (95% CI) for each subgroup of participants. The baseline visit (month 0 of follow-up) corresponds to 24 months after antiretroviral therapy initiation. The OR of presenting a clinical event corresponds to the five-year follow-up period.

Subanalyses according to event type yielded consistent estimates for all outcomes (MACE, NADM, or non-accidental death). However, they only reached statistical significance for death and MACE at a CD4/CD8 ratio cut-off of <0.2. The CD4/CD8 ratio was more predictive of death or MACE than NADM (OR 3.52, 95% CI 1.03 - 12.4; OR 4.4, 95% CI 1.4 - 12.7, and OR 2.4, 95% CI 0.8 - 6.9, respectively) (**Supplementary Table S1**). The *on-treatment* secondary analysis showed results similar to those of the main analysis (**Supplementary Table S2**).

The CD4/CD8 ratio correlated with both the CD4 count (Rho 0.53, p < 0.001) and CD8 count (Rho -0.47, p < 0.001). However, among patients with a CD4 count ≥500 cells/μL, the correlation with the CD4 count became weaker (Rho 0.31, p < 0.001), while that with the CD8 count was stronger (Rho -0.63, p < 0.001).

## Discussion

In this prospective multicenter cohort of treatment-naïve PWH, we compared the ability of CD4 count, CD8 count, and CD4/CD8 ratio measured two years after ART to predict the risk of SNAEs and mortality over the subsequent five years. The ratio (<0.3) was the only variable that predicted the risk across the entire range of CD4 counts.

Was the association of the CD4/CD8 ratio with the risk of SNAE driven by CD4 count, CD8 count, or both? Interestingly, both the CD4 and CD8 count contributed to the risk prediction, but the extent of immune suppression influenced their contribution. In patients with a CD4 count <500/μL, the association between a low CD4/CD8 ratio and SNAE risk is mainly driven by CD4 count. Conversely, in patients with a higher CD4 count, the association depends on the CD8 count, consistent with our previous findings in a different cohort [8]. These results suggest that, in patients with a low CD4 count, much of the risk associated with a low ratio is driven by immunodeficiency. In contrast, the risk associated with a low ratio in those with a higher CD4 count is driven by inflammation and immunosenescence (reflected as a high CD8 count). Both mechanisms seem to be captured by the CD4/CD8 ratio across the entire CD4 spectrum, although the CD4/CD8 ratio predictive capacity becomes stronger as the CD4 count increases.

The most predictive CD4/CD8 ratio and CD8 count cut-off values represent an area of controversy. In this study, the most extreme CD4 count, CD8 count, and CD4/CD8 ratio cut-off values (<200 cells/μL, ≥1500 cells/μL, and <0.2, respectively) predicted the most significant SNAEs risk. However, the association disappeared at values far outside their normal range in the general population (≥500 cells/μL, <1000 cells/μL, and ≥0.5, respectively). While a CD4/CD8 ratio <1 is considered abnormal in the general population [22] and an independent predictor of mortality [23,24], the finding that the CD4/CD8 ratio cut-off for predicting SNAEs risk in PWH on ART is much lower (0.4) is consistent with some previous studies [8,9,25,26]. For CD8 count, to the best of our knowledge, no studies have investigated the most discriminative cut-off values. Consistent with our findings, two previous studies have reported that a CD8 count >1500 cells/μL predicted SNAEs. In the Copenhagen cohort, a CD8 count >1500 cells/μL measured after ten years of ART predicted an 80% increased risk of non-AIDS-associated mortality compared to a lower CD8 count [19]. In the AIDS clinical trials group longitudinal linked randomized trials (ALLRT) cohort, a CD8 count >1500 cells/μL at year 2 of ART predicted a 75% increased risk of AIDS and non-infectious non-AIDS events during the following 5 years of treatment [14].

In addition to the different CD4/CD8 ratios and CD8 counts used in previous literature to define associations with SNAEs risk, another important source of heterogeneity across studies is the timing of CD4 and CD8 measurements relative to ART initiation, including time-varying variables [11,12,15,27,28] These variables are less interpretable and more challenging to translate to the clinic [29]. Furthermore, measurements taken proximal to the event can make the role of the CD4/CD8 as a predictor less clear [13,25,30]. Instead, we used a landmark analysis, that is, we measured our predictor variable at year two from ART initiation. Landmark analysis offers several advantages: (i) Simplicity: the results are more interpretable for clinicians. Our analysis provides information on severe non-AIDS events predicted by the CD4/CD8 ratio or CD8 lymphocytes measured at a time when most immune activation markers are in a plateau phase [31,32]; (ii) Methodological: a time-updated CD4/CD8 ratio would be affected by the CD4 drop known to occur before the development of severe non-AIDS events [33]; iii) Consistency and reproducibility with subsequent work: landmark analysis facilitates comparison of the effects across studies [14,34,35]; and iv) Time horizon, as this strategy allowed us to predict the mid-term risk of clinical progression (through years 3-7), which will be more informative for clinicians.

The major strength of this study is that, in contrast to other studies, the statistical methodology we followed allowed us to simultaneously control for confounding and possible selection bias due to informative censoring, an issue that should be considered especially in cohort studies with prolonged follow-up. The use of adjusted survival curves overcomes the shortcomings of using the hazard ratio as a measure of effect, which can sometimes be uninformative [36]. In addition, we used a robust definition of SNAEs, which were prospectively adjudicated in CoRIS. Specifically, we included MACEs and NADMs, as these are the SNAEs that have demonstrated a more consistent association with the CD4/CD8 ratio in prior research [7,9–12,25].

However, certain limitations of this study must be considered. First, although we used a prospective multicenter cohort with a large sample size and long follow-up, as in any observational study, there is a risk of unmeasured confounding. In addition, this cohort has a small representation of women and certain ethnic groups, as well as INSTI-based first-line ART, the therapy currently recommended in clinical guidelines [16,17], contrary to when the study participants started ART.

We believe that the field needs to unify the criteria for CD4/CD8 and CD8 evaluation, and we propose using landmark analysis, as in this study. This could help in systematic review evaluations and allow external validation of the results. A future direction is to understand whether these altered immune profiles (low CD4/CD8 ratio or high CD8 count) protect against certain non-AIDS events, such as the risk of bacterial infections, as suggested in a previous study [14]. In addition, it is possible that the CD4/CD8 ratio and CD8 count are linked to some, but not all, types of non-AIDS events. In our study, their association with the risk of non-accidental mortality and MACE was stronger than that with the risk of NADM. Future studies should investigate which types of events are more strongly associated with the CD4/CD8 ratio and CD8 count, and explore the potential mechanisms.

In summary, the results of this large prospective cohort support the use of a CD4/CD8 ratio cut-off of <0.3, regardless of CD4 count, to identify PWH with an excess risk of cardiovascular events, non-AIDS-defining malignancies, and all-cause mortality. A CD8 count of ≥1500 cells/μL also predicts an increased risk, but the CD4 count does not add prognostic information beyond 500 cells/μL. In addition to identifying PWH that might benefit from closer monitoring, our results encourage the evaluation of the CD4/CD8 ratio and CD8 count as surrogate endpoints for new therapies for PWH.

## Data Availability

All data produced in the present study are available upon reasonable request to the authors

## Acknowledgments

We acknowledge all study participants who made this research possible.

## References

1. Edwards JK, Cole SR, Breger TL, et al. Mortality among people entering HIV care compared to the general US population: an observational study. Ann Intern Med 2021; 174:1197.

2. Edwards JK, Cole SR, Breger TL, et al. Five-Year Mortality for Adults Entering Human Immunodeficiency Virus Care Under Universal Early Treatment Compared With the General US Population. Clinical Infectious Diseases 2022; 75:867–874.

3. Trickey A, Sabin CA, Burkholder G, et al. Life expectancy after 2015 of adults with HIV on long-term antiretroviral therapy in Europe and North America: a collaborative analysis of cohort studies. Lancet HIV 2023; S2352-3018(23)00028-0 (Online ahead of print).

4. Sereti I, Krebs SJ, Phanuphak N, et al. Persistent, Albeit Reduced, Chronic Inflammation in Persons Starting Antiretroviral Therapy in Acute HIV Infection. Clin Infect Dis 2017; 64:124–131.

5. Lewden C, Bouteloup V, De Wit S, et al. All-cause mortality in treated HIV-infected adults with CD4 ≥500/mm3 compared with the general population: evidence from a large European observational cohort collaboration. Int J Epidemiol 2012; 41:433–445.

6. Borges ÁH, Committee for the IS and ESG and the SS, O’Connor JL, et al. Factors Associated With Plasma IL-6 Levels During HIV Infection. J Infect Dis 2015; 212:585–595

7. Ron R, Moreno E, Martínez-Sanz J, et al. CD4/CD8 ratio during HIV treatment: time for routine monitoring? Clinical Infectious Diseases 2023; ciad136 (Online ahead of print).

8. Serrano-Villar S, Sainz T, Lee S a., et al. HIV-Infected Individuals with Low CD4/CD8 Ratio despite Effective Antiretroviral Therapy Exhibit Altered T Cell Subsets, Heightened CD8+ T Cell Activation, and Increased Risk of Non-AIDS Morbidity and Mortality. PLoS Pathog 2014; 10:e1004078.

9. Mussini C, Lorenzini P, Cozzi-Lepri A, et al. CD4/CD8 ratio normalisation and non-AIDS-related events in individuals with HIV who achieve viral load suppression with antiretroviral therapy: An observational cohort study. Lancet HIV 2015; 2:98–106.

10. Castilho JL, Shepherd BE, Koethe J, et al. CD4+/CD8+ ratio, age, and risk of serious noncommunicable diseases in HIV-infected adults on antiretroviral therapy. AIDS 2016; 30:899–908.

11. Sigel K, Wisnivesky J, Crothers K, et al. Immunological and infectious risk factors for lung cancer in US veterans with HIV: a longitudinal cohort study. Lancet HIV 2017; 4:e67–e73.

12. Castilho JL, Bian A, Jenkins CA, et al. CD4/CD8 Ratio and Cancer Risk Among Adults With HIV. J Natl Cancer Inst 2022; 114:854–862.

13. Novak RM, Armon C, Battalora L, et al. Aging, trends in CD4+/CD8+ cell ratio, and clinical outcomes with persistent HIV suppression in a dynamic cohort of ambulatory HIV patients. AIDS 2022; 36:815–827.

14. Serrano-Villar S, Wu K, Hunt PW, et al. Predictive value of CD8+ T cell and CD4/CD8 ratio at two years of successful ART in the risk of AIDS and non-AIDS events. EBioMedicine 2022; 80: 104072.

15. Trickey A, May MT, Schommers P, et al. CD4:CD8 Ratio and CD8 Count as Prognostic Markers for Mortality in Human Immunodeficiency Virus-Infected Patients on Antiretroviral Therapy: The Antiretroviral Therapy Cohort Collaboration (ART-CC). 2017; 65(6):959–966

16. Panel on Antiretroviral Guidelines for Adults and Adolescents. Guidelines for the Use of Antiretroviral Agents in Adults and Adolescents Living with HIV. Department of Health and Human Services. Available at http://www.aidsinfo.nih.gov/ContentFiles/Adult. Available at: https://aidsinfo.nih.gov/guidelines. Accessed 21 March 2023.

17. European AIDS Clinical Society. EACS Guidelines version 11.1, October 2022. Available at: https://www.eacsociety.org/guidelines/eacs-guidelines/. Accessed 21 March 2023.

18. Hema MN, Ferry T, Dupon M, et al. Low CD4/CD8 Ratio Is Associated with Non AIDS-Defining Cancers in Patients on Antiretroviral Therapy: ANRS CO8 (Aproco/Copilote) Prospective Cohort Study. PLoS One 2016; 11:e0161594.

19. Helleberg M, Kronborg G, Ullum H, Ryder LP, Obel N, Gerstoft J. Course and Clinical Significance of CD8+ T-Cell Counts in a Large Cohort of HIV-Infected Individuals. J Infect Dis 2015; 211:1726–34.

20. Sobrino-Vegas P, Gutiérrez F, Berenguer J, et al. The Cohort of the Spanish HIV Research Network (CoRIS) and its associated biobank; organizational issues, main findings and losses to follow-up. Enferm Infecc Microbiol Clin 2011; 29:645–53.

21. Cole SR, Herná N MA. Constructing Inverse Probability Weights for Marginal Structural Models. Am J Epidemiol 2008; 168(6):656–64.

22. Wikby A, Maxson P, Olsson J, Johansson B, Ferguson FG. Changes in CD8 and CD4 lymphocyte subsets, T cell proliferation responses and non-survival in the very old: the Swedish longitudinal OCTO-immune study. Mech Ageing Dev 1998; 102:187–198.

23. Olsson J, Wikby A, Johansson B, Löfgren S, Nilsson BO, Ferguson FG. Age-related change in peripheral blood T-lymphocyte subpopulations and cytomegalovirus infection in the very old: the Swedish longitudinal OCTO immune study. Mech Ageing Dev 2001; 121:187–201.

24. Ferguson FG, Wikby A, Maxson P, Olsson J, Johansson B. Immune Parameters in a Longitudinal Study of a Very Old Population of Swedish People: A Comparison Between Survivors and Nonsurvivors. The Journals of Gerontology: Series A 1995; 50A:B378–B382.

25. Serrano-Villar S, Pérez-Elías MJ, Dronda F, et al. Increased risk of serious non-AIDS-related events in HIV-infected subjects on antiretroviral therapy associated with a low CD4/CD8 ratio. PLoS One 2014; 9:e85798.

26. Han WM, Apornpong T, Kerr SJ, et al. CD4/CD8 ratio normalization rates and low ratio as prognostic marker for non-AIDS defining events among long-term virologically suppressed people living with HIV. AIDS Res Ther 2018; 15:13.

27. Domínguez-Domínguez L, Rava M, Bisbal O, et al. Low CD4/CD8 ratio is associated with increased morbidity and mortality in late and non-late presenters: results from a multicentre cohort study, 2004–2018. BMC Infect Dis 2022; 22:1–10.

28. Chammartin F, Darling K, Abela IA, et al. CD4:CD8 Ratio and CD8 Cell Count and Their Prognostic Relevance for Coronary Heart Disease Events and Stroke in Antiretroviral Treated Individuals: The Swiss HIV Cohort Study. J Acquir Immune Defic Syndr 2022; 91:508.

29. Fisher LD, Lin DY. Time-dependent covariates in the Cox proportional-hazards regression model. Annu Rev Public Health 2003; 20:145–157.

30. Castilho JL, Turner M, Shepherd BE, et al. CD4/CD8 Ratio and CD4 Nadir Predict Mortality Following Noncommunicable Disease Diagnosis in Adults Living with HIV. AIDS Res Hum Retroviruses 2019; 35:960.

31. Gandhi RT, Spritzler J, Chan E, et al. Effect of baseline- and treatment-related factors on immunologic recovery after initiation of antiretroviral therapy in HIV-1-positive subjects: Results from ACTG 384. J Acquir Immune Defic Syndr (1988) 2006; 42:426–434.

32. Serrano-Villar S, Martínez-Sanz J, Ron R, et al. Effects of first-line antiretroviral therapy on the CD4/CD8 ratio and CD8 cell counts in CoRIS: a prospective multicentre cohort study. Lancet HIV 2020; 7:e565–e573.

33. Helleberg M, Kronborg G, Larsen CS, et al. CD4 Decline Is Associated With Increased Risk of Cardiovascular Disease, Cancer, and Death in Virally Suppressed Patients With HIV. Clinical Infectious Diseases 2013; 57:314–321.

34. McComsey GA, Kitch D, Sax PE, et al. Associations of Inflammatory Markers with AIDS and non-AIDS Clinical Events after Initiation of Antiretroviral Therapy: AIDS Clinical Trials Group A5224s, a substudy of ACTG A5202. J Acquir Immune Defic Syndr 2014; 65:167.

35. Angelidou K, Hunt PW, Landay AL, et al. Changes in Inflammation but Not in T-Cell Activation Precede Non-AIDS-Defining Events in a Case-Control Study of Patients on Long-term Antiretroviral Therapy. J Infect Dis 2018; 218:239.

36. Hernán MA. The Hazards of Hazard Ratios. Epidemiology 2010; 21:13.

