## Supplementary material for "Expanding HIV Clinical Monitoring: The Role of CD4, CD8, and CD4/CD8 Ratio in Predicting Non-AIDS Events"

**Table S1. Subanalyses according to type of event**

| **Event and cut-off** | **OR (95% CI)** | ***p*** |
| --- | --- | --- |
| **Nonaccidental death (n = 47)** |  |  |
| CD4/CD8 ratio < 0.2 | 3.58 (1.03, 12.36) | 0.043 |
| CD4/CD8 ratio < 0.3 | 1.40 (0.50, 3.91) | 0.525 |
| CD4/CD8 ratio < 0.4 | 1.00 (0.39, 2.50) | 0.988 |
| CD4/CD8 ratio < 0.5 | 1.06 (0.47, 2.40) | 0.881 |
| **MACE (n = 48)** |  |  |
| CD4/CD8 ratio < 0.2 | 4.14 (1.35, 12.66) | 0.013 |
| CD4/CD8 ratio < 0.3 | 1.52 (0.66, 3.50) | 0.324 |
| CD4/CD8 ratio < 0.4 | 1.10 (0.50, 2.43) | 0.817 |
| CD4/CD8 ratio < 0.5 | 1.75 (0.87, 3.51) | 0.113 |
| **NADM (n = 105)** |  |  |
| CD4/CD8 ratio < 0.2 | 2.40 (0.83, 6.91) | 0.107 |
| CD4/CD8 ratio < 0.3 | 1.49 (0.81, 2.72) | 0.201 |
| CD4/CD8 ratio < 0.4 | 1.48 (0.83, 2.65) | 0.181 |
| CD4/CD8 ratio < 0.5 | 1.41 (0.84, 2.38) | 0.189 |

*Abbreviations: MACE, major adverse cardiovascular event; NADM, non-AIDS-defining malignancy*

**Table S2. On-treatment analysis**

| **Cut-off at year 2** | **OR (95% CI)** | ***p*** |
| --- | --- | --- |
| **CD4/CD8 ratio** |  |  |
| ≥ 0.2 | 0.32 (0.16, 0.64) | 0.001 |
| ≥ 0.3 | 0.57 (0.36, 0.91) | 0.019 |
| ≥ 0.4 | 0.69 (0.45, 1.05) | 0.090 |
| ≥ 0.5 | 0.75 (0.52, 1.11) | 0.143 |
| **CD4+ count (cells/μL)** |  |  |
| ≥ 200 | 0.26 (0.11, 0.57) | 0.001 |
| ≥ 350 | 0.35 (0.22, 0.56) | <0.001 |
| ≥ 500 | 0.58 (0.39, 0.87) | 0.008 |
| ≥ 750 | 0.73 (0.45, 1.18) | 0.197 |

*For the on-treatment analysis, we assumed that no participant discontinued antiretroviral treatment during follow-up. Only censoring due to loss to follow-up was considered for inverse probability weighting.*

**Supplementary Figure S1.**

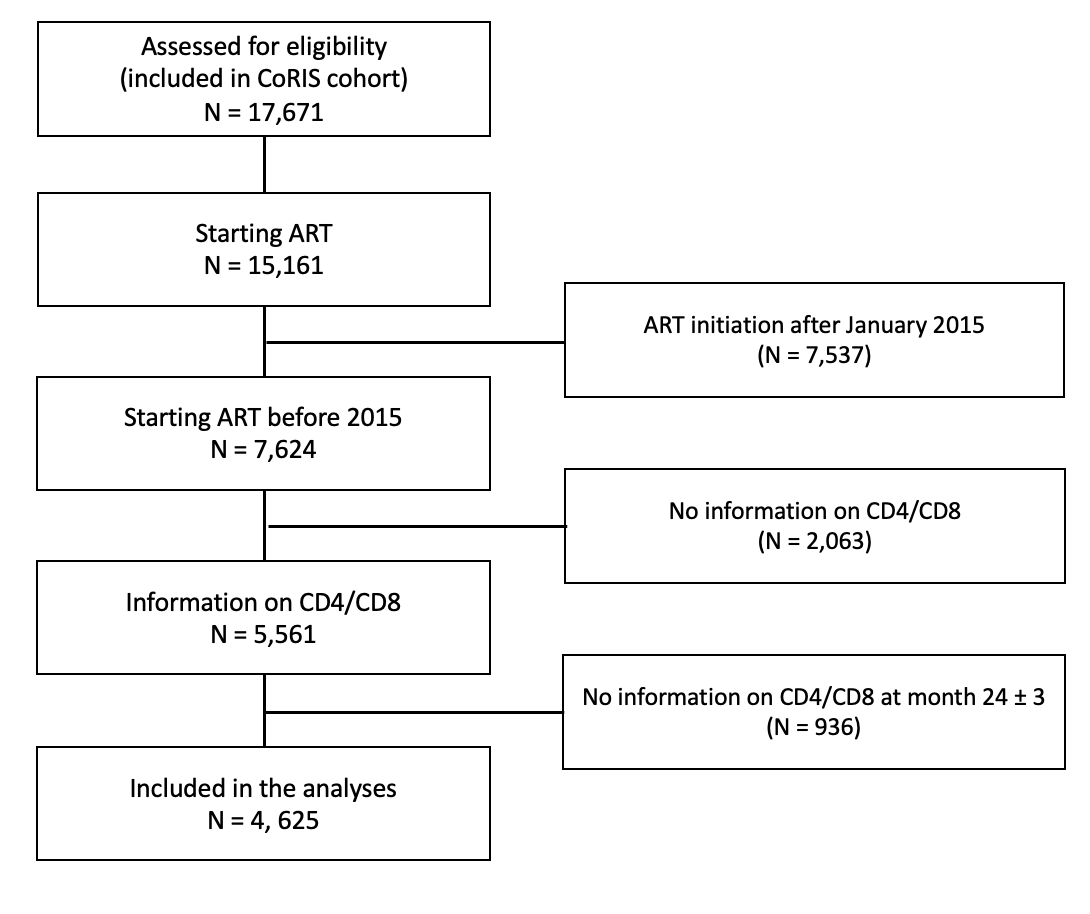

**Study flowchart**

**Supplementary Figure S2**

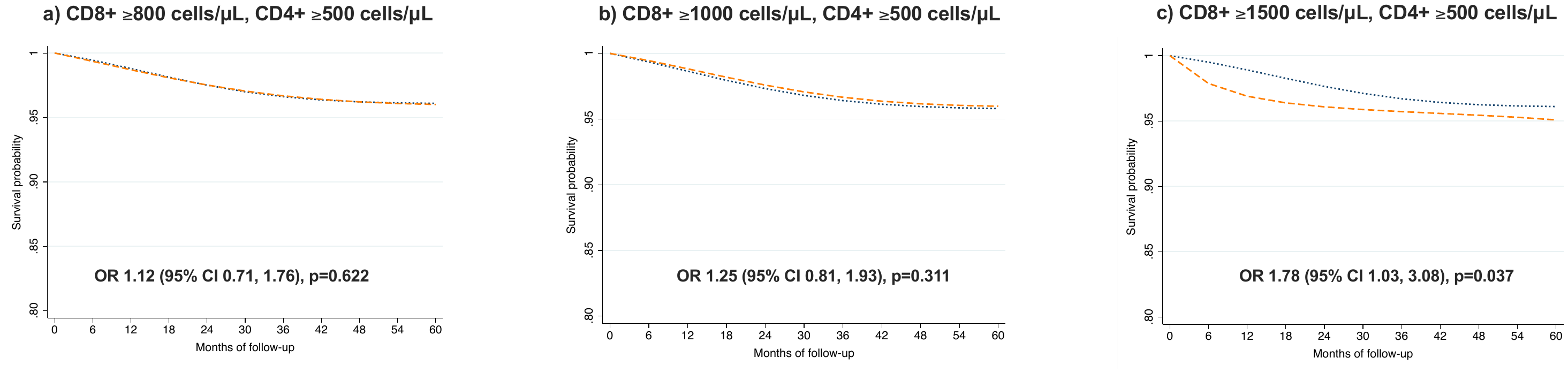

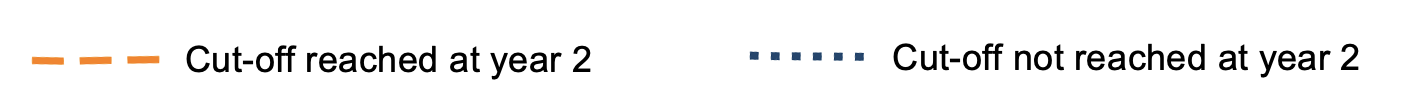

**Survival curves for each CD8+ cut-off among participants with CD4+ count ≥ 500 cells/μL.**

Survival probability and odds ratio (95% CI) for each subgroup of participants. The baseline visit (month 0 of follow-up) corresponds to 24 months after antiretroviral therapy initiation. The OR of presenting a clinical event corresponds to the five-year follow-up period.

**CENTERS AND RESEARCHERS INVOLVED IN CoRIS**

**Executive committee**

Santiago Moreno, Inma Jarrín, David Dalmau, M Luisa Navarro, M Isabel González, Federico Garcia, Eva Poveda, Jose Antonio Iribarren, Félix Gutiérrez, Rafael Rubio, Francesc Vidal, Juan Berenguer, Juan González, M Ángeles Muñoz-Fernández.

**Fieldwork data management and analysis**

Inmaculada Jarrín, Cristina Moreno, Marta Rava, Rebeca Izquierdo.

**BioBanK HIV Hospital General Universitario Gregorio Marañón**

M Ángeles Muñoz-Fernández, Elba Mauleón.

**Hospital General Universitario de Alicante (Alicante)**

Joaquín Portilla, Irene Portilla, Esperanza Merino, Gema García, Iván Agea, José Sánchez-Payá, Juan Carlos Rodríguez, Livia Giner, Sergio Reus, Vicente Boix, Diego Torrus, Verónica Pérez, Julia Portilla.

**Hospital Universitario de Canarias (San Cristóbal de la Laguna)**

Juan Luís Gómez, Jehovana Hernández, Ana López Lirola, Dácil García, Felicitas Díaz-Flores, M Mar Alonso, Ricardo Pelazas, M Remedios Alemán.

**Hospital Universitario Central de Asturias (Oviedo)**

Víctor Asensi, María Eugenia Rivas Carmenado, Tomás Suarez-Zarracina.

**Hospital Universitario 12 de Octubre (Madrid)**

Federico Pulido, Rafael Rubio, Otilia Bisbal, M Asunción Hernando, David Rial, María de Lagarde, Octavio Arce, Adriana Pinto, Laura Bermejo, Mireia Santacreu, Roser Navarro, Candela Gonzalez.

**Servicio de Enfermedades Infecciosas. Hospital Universitario Donostia. Instituto de Investigación BioDonostia (Donostia-San Sebastián)**

Jose Antonio Iribarren, M José Aramburu, Xabier Camino, Miguel Ángel von Wichmann, Miguel Ángel Goenaga, M Jesús Bustinduy, Harkaitz Azkune, Maialen Ibarguren, Xabier Kortajarena, Ignacio Álvarez-Rodriguez, Leire Gil, Lourdes Martínez.

**Hospital General Universitario De Elche (Elche)**

Félix Gutiérrez, Catalina Robledano, Mar Masiá, Sergio Padilla, Araceli Adsuar, Rafael Pascual, Marta Fernández, Antonio Galiana, José Alberto García, Xavier Barber, Vanessa Agullo, Javier Garcia Abellán, Reyes Pascual, Guillermo Telenti, Lucia Guillén, Ángela Botella.

**Hospital Universitari Germans Trias i Pujol (Can Ruti) (Badalona)**

Roberto Muga, Arantza Sanvisens, Daniel Fuster.

**Hospital General Universitario Gregorio Marañón (Madrid)**

Juan Berenguer, Isabel Gutierrez, Juan Carlos López, Margarita Ramírez, Belén Padilla, Paloma Gijón, Teresa Aldamiz-Echevarría, Francisco Tejerina, Cristina Diez, Leire Pérez, Chiara Fanciulli, Saray Corral.

**Hospital Universitari de Tarragona Joan XXIII (Tarragona)**

Francesc Vidal, Anna Martí, Joaquín Peraire, Consuelo Viladés, Montserrat Vargas, Montserrat Olona, Anna Rull, Verónica Alba, Elena Yeregui, Jenifer Masip, Graciano García-Pardo, Frederic Gómez Bertomeu, Sonia Espineira.

**Hospital Universitario y Politécnico de La Fe (Valencia)**

Marta Montero, Sandra Cuéllar, Marino Blanes, María Tasias, Eva Calabuig, Miguel Salavert, Juan Fernández, Inmaculada Segarra.

**Hospital Universitario La Paz/IdiPAZ**

Juan González-García, Ana Delgado, Francisco Arnalich, José Ramón Arribas, Jose Ignacio Bernardino, Juan Miguel Castro, Luis Escosa, Pedro Herranz, Victor Hontañón, Silvia García-Bujalance, Milagros García, Alicia González-Baeza, M Luz Martín-Carbonero, Mario Mayoral, M Jose Mellado, Rafael Esteban, Rocío Montejano, M Luisa Montes, Victoria Moreno, Ignacio Pérez-Valero, Berta Rodés, Guadalupe Rúa, Talía Sainz, Elena Sendagorta, Eulalia Valencia, Carmen Busca, Joanna Cano, Julen Cardiñanos, Rosa de Miguel.

**Hospital San Pedro Centro de Investigación Biomédica de La Rioja (CIBIR) (Logroño)**

Jose Ramón Blanco, Laura Pérez-Martínez, José Antonio Oteo, Valvanera Ibarra, Luis Metola, Mercedes Sanz.

**Hospital Universitario Miguel Servet (Zaragoza)**

Piedad Arazo, Gloria Sampériz.

**Hospital Universitari Mutua Terrassa (Terrassa)**

David Dalmau, Marina Martinez, Angels Jaén, Montse Sanmartí, Mireia Cairó, Javier Martinez-Lacasa, Pablo Velli, Roser Font, Mariona Xercavins, Noemí Alonso, Francesco Aiello.

**Complejo Hospitalario de Navarra (Pamplona)**

María Rivero, Beatriz Piérola, Maider Goikoetxea, María Gracia, Carlos Ibero, Estela Moreno, Jesús Repáraz.

**Parc Taulí Hospital Universitari (Sabadell)**

Gemma Navarro, Manel Cervantes Garcia, Sonia Calzado Isbert, Marta Navarro Vilasaro, Belen Lopez Garcia.

**Hospital Universitario de La Princesa (Madrid)**

Ignacio de los Santos, Alejandro de los Santos, Jesús Sanz, Lucio García-Fraile, Enrique Martín, Ildefonso Sánchez-Cerrillo, Marta Calvet, Ana Barrios, Azucena Bautista, Carmen Sáez, Marianela Ciudad, Ángela Gutiérrez.

**Hospital Universitario Ramón y Cajal (Madrid)**

Santiago Moreno, Santos del Campo, José Luis Casado, Fernando Dronda, Ana Moreno, M Jesús Pérez, Sergio Serrano, Mª Jesús Vivancos, Javier Martínez-Sanz, Alejandro Vallejo, Matilde Sanchez, Jose Antonio Pérez-Molina, José Manuel Hermida.

**Hospital General Universitario Reina Sofía (Murcia)**

Enrique Bernal, Antonia Alcaraz, Joaquín Bravo, Ángeles Muñoz, Cristina Tomás, Mónica Martínez, M Carmen Villalba.

**Hospital Nuevo San Cecilio (Granada)**

Federico García, Clara Martínez, José Hernández, Leopoldo Muñoz Medina, Marta Álvarez, Natalia Chueca, David Vinuesa, Adolfo de Salazar, Ana Fuentes, Emilio Guirao, Laura Viñuela, Andrés Ruiz-Sancho, Francisco Anguita.

**Centro Sanitario Sandoval (Madrid)**

Jorge Del Romero, Montserrat Raposo, Carmen Rodríguez, Teresa Puerta, Juan Carlos Carrió, Mar Vera, Juan Ballesteros, Oskar Ayerdi, Begoña Baza, Eva Orviz.

**Hospital Clínico Universitario de Santiago (Santiago de Compostela)**

Antonio Antela, Elena Losada.

**Hospital Universitario Son Espases (Palma de Mallorca)**

Melchor Riera, María Peñaranda, M Angels Ribas, Antoni A. Campins, Mercedes Garcia-Gazalla, Francisco J Fanjul, Javier Murillas, Francisco Homar, Helem H Vilchez, Luisa Martin, Antoni Payeras.

**Hospital Universitario Virgen de la Victoria (Málaga)**

Jesús Santos, María López, Crisitina Gómez, Isabel Viciana, Rosario Palacios.

**Hospital Universitario Virgen del Rocío (Sevilla)**

Luis Fernando López-Cortés, Nuria Espinosa, Cristina Roca, Silvia Llaves.

**Hospital Universitario de Bellvitge (Hospitalet de Llobregat)**

Juan Manuel Tiraboschi, Arkaitz Imaz, Ana Karina Silva, María Saumoy, Sofía Catalina Scévola.

**Hospital Universitario Valle de Hebrón (Barcelona)**

Adrián Curran, Vicenç Falcó, Jordi Navarro, Joaquin Burgos, Paula Suanzes, Jorge García, Vicente Descalzo, Patricia Álvarez, Bibiana Planas, Marta Sanchiz, Lucía Rodríguez.

**Hospital Costa del Sol (Marbella)**

Julián Olalla, M José Sánchez, Javier Pérez, Alfonso del Arco, Javier de la Torre, José Luis Prada.

**Hospital General Universitario Santa Lucía (Cartagena)**

Onofre Juan Martínez, Lorena Martinez, Francisco Jesús Vera, Josefina García, Begoña Alcaraz, Antonio Jesús Sánchez Guirao .

**Complejo Hospitalario Universitario a Coruña (Chuac) (A Coruña)**

Alvaro Mena, Angeles Castro, Berta Pernas, Pilar Vázquez, Soledad López.

**Hospital Universitario Basurto (Bilbao)**

Sofía Ibarra, Guillermo García, Josu Mirena, Oscar Luis Ferrero, Josefina López, M Mar Cámara, Mireia de la Peña, Miriam Lopez, Iñigo Lopez, Itxaso Lombide, Victor Polo, Joana de Miguel.

**Hospital Universitario Virgen de la Arrixaca (El Palmar)**

Carlos Galera, Marian Fernández, Helena Albendin, Antonia Castillo, Asunción Iborra, Antonio Moreno, M Angustias Merlos, Asunción Vidal.

**Hospital de la Marina Baixa (La Vila Joiosa)**

Concha Amador, Francisco Pasquau, Concepcion Gil, Jose Tomás Algado.

**Hospital Universitario Infanta Sofía (San Sebastián de los Reyes)**

Inés Suarez-García, Eduardo Malmierca, Patricia González-Ruano, M Pilar Ruiz, José Francisco Pascual, Elena Sáez, Luz Balsalobre.

**Hospital Universitario de Jaén (Jaén)**

M Villa López, Mohamed Omar, Carmen Herrero, M Amparo Gómez.

**Hospital Universitario San Agustín (Avilés)**

Miguel Alberto de Zarraga, Desiré Pérez.

**Hospital Clínico San Carlos (Madrid)**

Vicente Estrada, Nieves Sanz, Noemí Cabello, Jorge Vergas García, Maria Jose Núñez, Iñigo Sagastagoitia.

**Hospital Universitario Fundación Jiménez Díaz (Madrid)**

Miguel Górgolas, Alfonso Cabello, Beatriz Álvarez, Laura Prieto, Irene Carrillo.

**Hospital Universitario Príncipe de Asturias (Alcalá de Henares)**

José Sanz, Alberto Arranz, Cristina Hernández, María Novella.

**Hospital Clínico Universitario de Valencia (Valencia)**

M José Galindo, Ana Ferrer.

**Hospital Reina Sofía (Córdoba)**

Antonio Rivero Román, Inma Ruíz, Antonio Rivero Juárez, Pedro López, Isabel Machuca, Mario Frias, Ángela Camacho, Ignacio Pérez, Diana Corona, Ignacio Pérez, Diana Corona.

**Hospital Universitario Severo Ochoa (Leganés)**

Miguel Cervero, Rafael Torres.

**Nuestra Señora de Valme (Sevilla)**

Juan Antonio Pineda, Pilar Rincón, Juan Macías, Luis Miguel Real, Anais Corma, Marta Fernández, Alejandro Gonzalez-Serna.

**Hospital Álvaro Cunqueiro (Vigo)**

Eva Poveda, Alexandre Pérez, Luis Morano, Celia Miralles, Antonio Ocampo, Guillermo Pousada, Lucía Patiño.

**Hospital Clínico Universitario de Valladolid (Valladolid)**

Carlos Dueñas, Sara Gutiérrez, Elena Tapia, Cristina Novoa, Xjoylin Egües, Pablo Telleria.
